# Cumulative Genetic Risk for Asthma Contributes to Disease Severity in Children with Asthma

**DOI:** 10.1101/2025.09.08.25335346

**Authors:** Matthew Dapas, William Wentworth-Sheilds, Emma E. Thompson, Rajesh Kumar, Elizabeth Lippner, Robert A. Wood, George T. O’Connor, Gurjit K. Khurana Hershey, Rebecca S. Gruchalla, Andrew H. Liu, Edward M. Zoratti, Leonard B. Bacharier, Stephanie Lovinsky-Desir, Michele A. Gill, William J. Sheehan, Shilpa J. Patel, Matthew C. Altman, James E. Gern, Cynthia M. Visness, Peter J. Gergen, Patrice M. Becker, Daniel J. Jackson, Carole Ober

## Abstract

**Background:** Childhood-onset asthma is highly heritable, with nearly 200 risk loci identified in genome-wide association studies. Aggregated polygenic risk scores can be used to quantify genetic predisposition to asthma, but their power to predict asthma severity in multi-ancestral groups has not been previously evaluated.

**Objective:** Our aim was to examine the predictive power of biobank-derived asthma polygenic risk scores in four multi-ancestry asthma study cohorts of children living in U.S. urban environments.

**Methods:** We generated polygenic risk scores for asthma, derived from a large-scale genome-wide association meta-analysis, and assessed genetic predictions across different subphenotypes of asthma and tested for associations between genetic asthma risk and measures of asthma severity.

**Results:** Polygenic risk score prediction was significantly stronger for more symptomatic asthma phenotypes (P<0.001), and scores were significantly higher in difficult-to-control vs. easy-to-control asthma (P=0.02). Genetic risk was also significantly associated with more frequent exacerbations (P=0.03), higher blood eosinophil levels (P=0.01), and lower lung function (P<0.001).

**Conclusion:** Cumulative genetic risk for asthma is associated with disease severity and exacerbation risk in children with asthma.

**Key Messages:**

- Polygenic risk prediction is stronger in more symptomatic phenotypes
- Cumulative genetic risk for asthma is associated with greater asthma severity, higher exacerbation frequency, lower lung function, and increased eosinophil levels in children with asthma

**Capsule Summary:** We demonstrate that cumulative genetic risk for asthma is associated with disease severity in children with asthma living in urban environments.

## INTRODUCTION

Asthma is a heterogenous disease with multiple endotypes and phenotypes, with severe forms of asthma accounting for a disproportionate share of the disease’s morbidity, mortality, and healthcare costs^1^. Identifying factors that contribute to disease severity could help improve risk stratification and guide personalized management strategies. Genetics contributes significantly to the overall risk of developing asthma, as evidenced by nearly 200 risk loci uncovered in large genome-wide association studies (GWASs)^2–5^. However, the relationship between cumulative genetic asthma risk and disease severity has not been fully explored^6^.

Genetic risk can be modeled using aggregated polygenic risk scores (PRSs). Many studies have developed predictive PRSs for asthma^7^, and phenome-wide association studies with asthma PRSs have suggested that genetic risk may correlate with asthma severity^6,7^. To directly test this relationship, we generated PRSs for asthma in four multi-ancestry asthma study cohorts of children living in low-income, urban environments, who face an excess burden of asthma morbidity and mortality^8–10^. We then assessed genetic predictions for different subphenotypes of asthma and tested for associations between genetic asthma risk and multiple measures of asthma severity.

## METHODS

### Cohorts

We analyzed samples and phenotypes from several National Institutes of Allergy and Infectious Diseases (NIAID)-funded asthma studies conducted by the Inner-City Asthma Consortium (ICAC)^11^: the Asthma Phenotypes in the Inner City (APIC) study^12,13^, the Urban Environment and Childhood Asthma (URECA) birth cohort study^14^, and the Mechanisms Underlying Asthma Exacerbations Prevented and Persistent with Immune Based Therapy: A Systems Approach (MUPPITS 1 & 2) studies^8,15^. Non-asthma, population-based controls from the Consortium on Asthma among African-ancestry Populations in the Americas (CAAPA) study^16^ was used as a comparator group to assess the distribution of the PRSs in an unselected healthy population with similar ancestry to the APIC, URECA, and MUPPITS participants. Ethical approval for this work was granted by a central institutional review board, WGC IRB^17^.

#### APIC

APIC was a prospective, observational study in children with asthma age 6-17 years old from low-income areas (≥20% of residents below federal poverty level) of nine U.S. cities. Hierarchical clustering was performed on a large collection of baseline demographic, clinical, and environmental variables to identify different phenotypes of childhood-onset asthma. Previously, five clusters were described (clusters A-E) based on varying symptoms, biomarkers, and disease severity^13^. APIC participants were also previously grouped based on required asthma controller treatment levels. Easy-to-control asthma was defined as requiring ≤100mcg/day of fluticasone at a majority of study visits, whereas difficult-to-control asthma was defined as requiring ≥500mcg/day^12^. APIC participants were genotyped using whole-genome sequencing (WGS) as previously described^18^.

#### URECA

URECA is a prospective, observational study of a birth cohort enriched for children with at least one parent with asthma or allergic disease in four U.S. cities^14^. All families lived in neighborhoods where at least 20% of the population had incomes below the poverty level. Clinical measurements at age 10 were used when available; otherwise, the most recent measurement after age 5 was used. URECA participants were genotyped using WGS as previously described^18^.

#### MUPPITS 1 & 2

The MUPPITS studies investigated exacerbations in children with exacerbation-prone eosinophilic asthma who were living in urban environments across nine U.S. cities. Participants in MUPPITS were 6-17 years old, had blood eosinophils of ≥150 cells/μL, required at least twice-daily treatment with inhaled corticosteroids, and had at least two exacerbations treated with systemic corticosteroids in the previous year. All participants lived in census tracts where at least 10% of families had incomes below the poverty level^19^. The MUPPITS-1 study was a 6-month prospective, longitudinal study that compared transcriptome profiles in children who experienced cold symptoms between those that subsequently experienced an exacerbation and those that did not^15^. The MUPPITS-2 study was a randomized controlled trial to assess the safety and efficacy of mepolizumab for reducing exacerbations in this population^8^. Genotypes for MUPPITS 1 & 2 were determined in 461 subjects with available DNA using the Illumina Genome Diversity Array (GDA, 8v1.0, hg19) at the University of Chicago. Individuals were removed if they met any of the following quality control (QC) criteria: genotyped vs. self-reported sex mismatches, unexpected sample duplication, >2% missing genotypes across all variants, or autosomal heterozygosity >5 standard deviations from the mean. Variants were excluded under the the following QC criteria: >4% missing genotypes across all samples or exact Hardy-Weinberg p-value <5×10^−8^. A total of 1,661,770 directly genotyped single-nucleotide variants remained following QC. Variants were lifted from human reference hg19 to hg38 using CrossMap^20^. Eagle^21^ was used for genotype phasing and minimac4^22^ was used for genotype imputation. Imputation was performed using the TOPMed reference panel (r2 version 1.0.0) of >100,000 whole genome sequences (human genome build 38) and the Michigan Imputation Server^23^. Following QC, 428 samples from MUPPITS 1 & 2 remained for analysis.

#### CAAPA

CAAPA^16^ performed WGS on approximately 1,000 individuals with African ancestry from 19 populations across the Americas and Caribbean as previously described^24^. The WGS data is available through dbGAP (accession phs001123.v2.p1). In this study, we utilized WGS data from CAAPA non-asthma control samples from among the African American populations (n=205). Variant QC followed the same criteria as applied to APIC and URECA WGS data^18^.

### Modeling Genetic Asthma Risk

Polygenic risk for asthma was modeled on published association summary statistics from a large-scale, multi-ancestry GWAS conducted as part of the Global Biobank Meta-analysis Initiative (GBMI)^2^. Summary statistics were available via https://www.globalbiobankmeta.org. The GWAS was a meta-analysis across 18 biobanks, consisting of 153,763 cases and 1,647,022 controls. Primary ancestry among asthma cases was 79.3% European, 12.1% East Asian, 3.3% African, 2.6% Admixed American, 2.6% Central and Southern Asian, and 0.01% Middle Eastern^2^. Asthma case and control statuses were assigned primarily according to “PheCodes” mapped from International Classification of Disease (ICD) codes, but several biobanks used self-reported data.

To generate individual PRSs, we applied the polygenic risk model published by Wang and colleagues^25^ from the GBMI multi-ancestry GWAS, which was generated using PRS-CS^26^ and deposited in the Polygenic Score Catalog (PGS001782)^27^. PRS-CS applies a Bayesian regression framework with continuous shrinkage priors on variant effect sizes from all tested variants. The “PRS-CS-auto” algorithm was used with default parameters to estimate the global and local shrinkage parameters directly from the GBMI GWAS summary statistics for each autosomal chromosome. Linkage disequilibrium was modeled using the EUR reference panel constructed by the PRS-CS developers with 1000 Genomes (1KG) Project data^25^. We aggregated the posterior variant effects into per-individual scores across the intersection of genotyped single-nucleotide variants present in all genotyped cohorts using the Plink (v2.00a5.8LM AVX2 Intel, 21 Nov 2023) “score” function^28^. Missing genotypes in individuals were imputed as the product of the variant’s posterior effect size and its effect-allele frequency^29^. Cumulative PRSs were converted to Z scores (across all genotyped cohorts) for analysis.

### Ancestry estimation

Principal components (PCs) of genetic ancestry were used to control for population stratification. Ancestry PCs were calculated on the intersect of high-quality single-nucleotide variants genotyped in the asthma cohorts and population reference panels from the 1KG Project (n=156)^30^ and the Human Genome Diversity (HGD) Project (n=52)^31^, as in our previous study^18^. Ancestry PCs were calculated accounting for subject relatedness using PC-Air^32^ and PC-Relate^33^, with initial kinship estimates derived using KING^34^. Kinship was estimated in iterations until resultant PCs stabilized (n=3).

### Association Testing

Differences in PRS distributions between cohorts were assessed using Wilcoxon rank-sum tests for pairwise comparisons and Kruskal-Wallis tests for more than two groups. PRS asthma prediction was conducted with the CAAPA non-asthma samples as controls. Statistical differences in receiver operating characteristic (ROC) areas under the curve (AUC) were evaluated using DeLong’s test. Differences in PRS distributions between difficult-to-control and easy-to-control treatment groups^12^ in APIC were assessed using logistic regression, adjusting for age, sex, and the first three ancestry PCs. PRS effects on exacerbation rates in MUPPITS-2 were determined using negative binomial generalized linear models, adjusting for age, sex, and the first three PCs of global ancestry. PRS associations with quantitative traits across multiple studies were assessed using a two-stage approach^18,35,36^. First, traits were regressed on age, sex, study, and the first three ancestry PCs. Then, model residuals were rank-normalized and regressed again on the same covariates with the addition of the PRS as a predictor. Adjusted trait values for plotting PRS associations were derived by adding first-stage model intercepts to residuals.

## RESULTS

We utilized genome-wide genotype data from four multi-ancestry asthma cohorts of children living in low-income, urban environments (APIC^13^, URECA^14^, MUPPITS-1^15^, and MUPPITS-2^8^), as summarized in **Table 1**, including 1,108 children with childhood-onset asthma. Nearly all of the participants in these studies were parent-identified as Black (66%) or Hispanic (26%), and the cohorts featured similar overall distributions of reported race/ethnicity (**Table 1**) and genetic ancestry (**Figure 1**).

**Figure 1.**
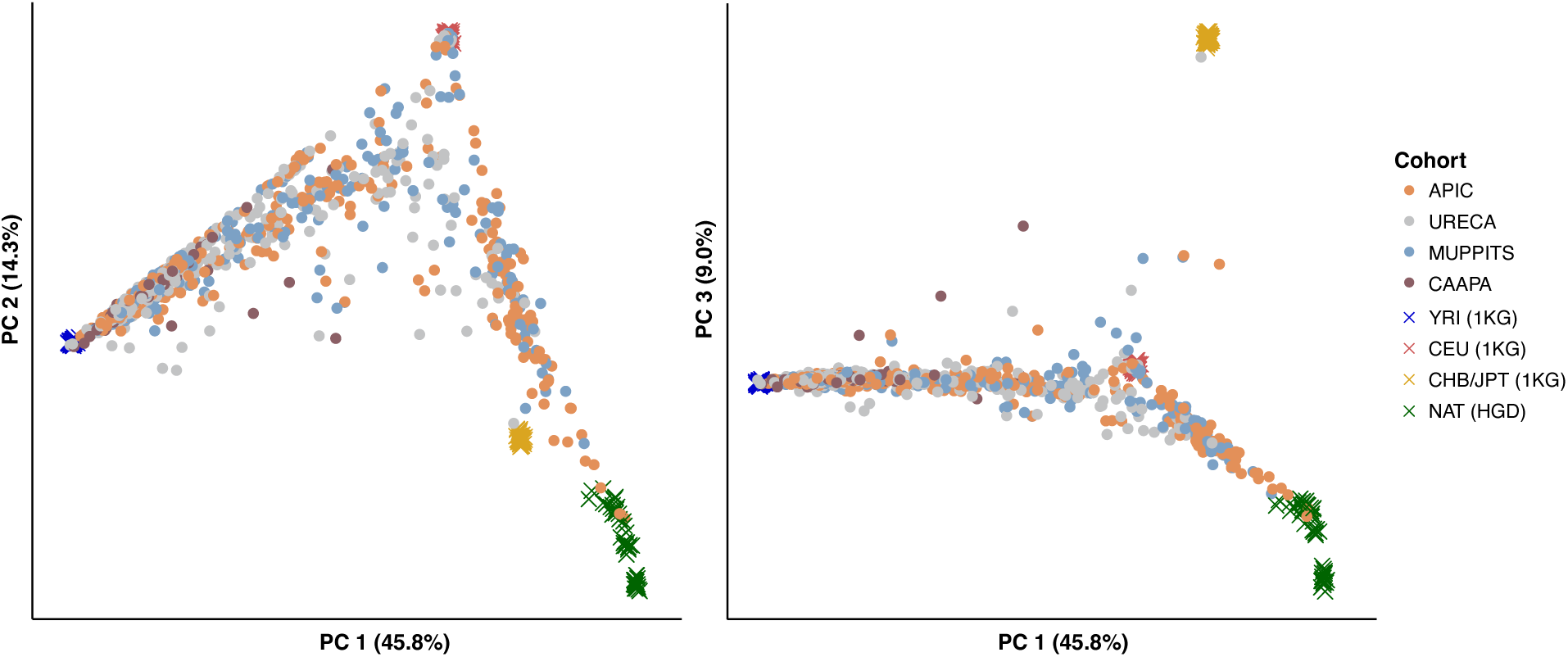
Principal components of ancestry by cohort. The top three principal components (PCs) of ancestry are plotted for sequenced APIC, URECA, MUPPITS, and CAAPA (non-asthma) participants, colored by study, along with the four ancestry reference populations used for determining ancestry. 1KG, 1000 Genomes Project; HGD, Human Genome Diversity Project.

**Table 1.** Demographic characteristics of childhood-onset asthma cohorts.

| Characteristic | All | APIC | URECA | MUPPITS 1 & 2 |
| --- | --- | --- | --- | --- |
| Number | 1462 | 507 | 527 | 428 |
| Age, years, median (range) | 10 (6-17) | 11 (6-17) | 10 (7-10) | 11 (6-17) |
| Female sex | 665 (45%) | 216 (43%) | 261 (50%) | 188 (44%) |
| *Race/Ethnicity* |  |  |  |  |
| Black (non-Hispanic) | 966 (66%) | 318 (63%) | 377 (72%) | 269 (63%) |
| White (non-Hispanic) | 23 (2%) | 7 (1%) | 7 (1%) | 9 (2%) |
| Hispanic | 378 (26%) | 154 (30%) | 104 (20%) | 120 (28%) |
| Other/mixed | 93 (6%) | 26 (5%) | 38 (7%) | 29 (7%) |
| Unknown | 4 (<1%) | 2 (<1%) | 1 (<1%) | 1 (<1%) |
| *Site* |  |  |  |  |
| Baltimore | 233 (16%) | 84 (17%) | 149 (28%) | - |
| Boston | 228 (16%) | 65 (13%) | 124 (24%) | 39 (9%) |
| Chicago | 97 (7%) | 62 (12%) | - | 35 (8%) |
| Cincinnati | 84 (6%) | 45 (9%) | - | 39 (9%) |
| Dallas | 86 (6%) | 38 (7%) | - | 48 (11%) |
| Denver | 103 (7%) | 59 (12%) | - | 44 (10%) |
| Detroit | 106 (7%) | 50 (10%) | - | 56 (13%) |
| New York | 227 (16%) | 65 (13%) | 99 (19%) | 63 (15%) |
| St. Louis | 207 (14%) | - | 155 (29%) | 52 (12%) |
| Washington, D.C | 91 (6%) | 39 (8%) | - | 52 (12%) |
| *Clinical traits* |  |  |  |  |
| Asthma Diagnosis | 1108 (75%) | 507 (100%) | 173 (33%) | 428 (100%) |
| Eosinophils/μL, median (IQR) | 300 (190-500) | 300 (160-485.5) | 200 (100-350) | 400 (300-600) |
| FEV<sub>1</sub>/FVC, median (IQR) | 0.80 (0.73-0.86) | 0.79 (0.72-0.85) | 0.84 (0.79-0.88) | 0.76 (0.68-0.82) |

APIC, Asthma Phenotypes in Inner-City Children; FEV<sub>1</sub>, Forced expiratory volume in one second; FVC, Forced vital capacity; IQR, interquartile range; MUPPITS, Mechanisms Underlying Asthma Exacerbations Prevented and Persistent with Immune Based Therapy: A Systems Approach; URECA, Urban Environment and Childhood Asthma

PRS distributions differed significantly by cohort (P<0.001, Kruskal-Wallis), with higher median PRSs observed in cohorts characterized by more severe forms of asthma (**Figures 2A, 2B**). Notably, the PRSs were significantly higher in URECA children without asthma, who were enriched for family history of asthma, than in population-based controls without asthma (CAAPA), likely owing to inherited disease risk (P=0.03, Wilcoxon). The PRS predicted asthma diagnosis more accurately in cohorts characterized by greater symptom burden, according to relative ROC AUCs (**Figure 2C**). Overall, the PRS yielded significantly better prediction for moderate-severe asthma (APIC clusters D and E, MUPPITS) (AUC=0.65) than for the other groups (AUC=0.53; P_Δ_=<0.001, **Figure 2D**). These results demonstrate that genetic asthma prediction is stronger for more symptomatic phenotypes and further suggest that genetic risk correlates with increasing disease severity in children.

**Figure 2.**
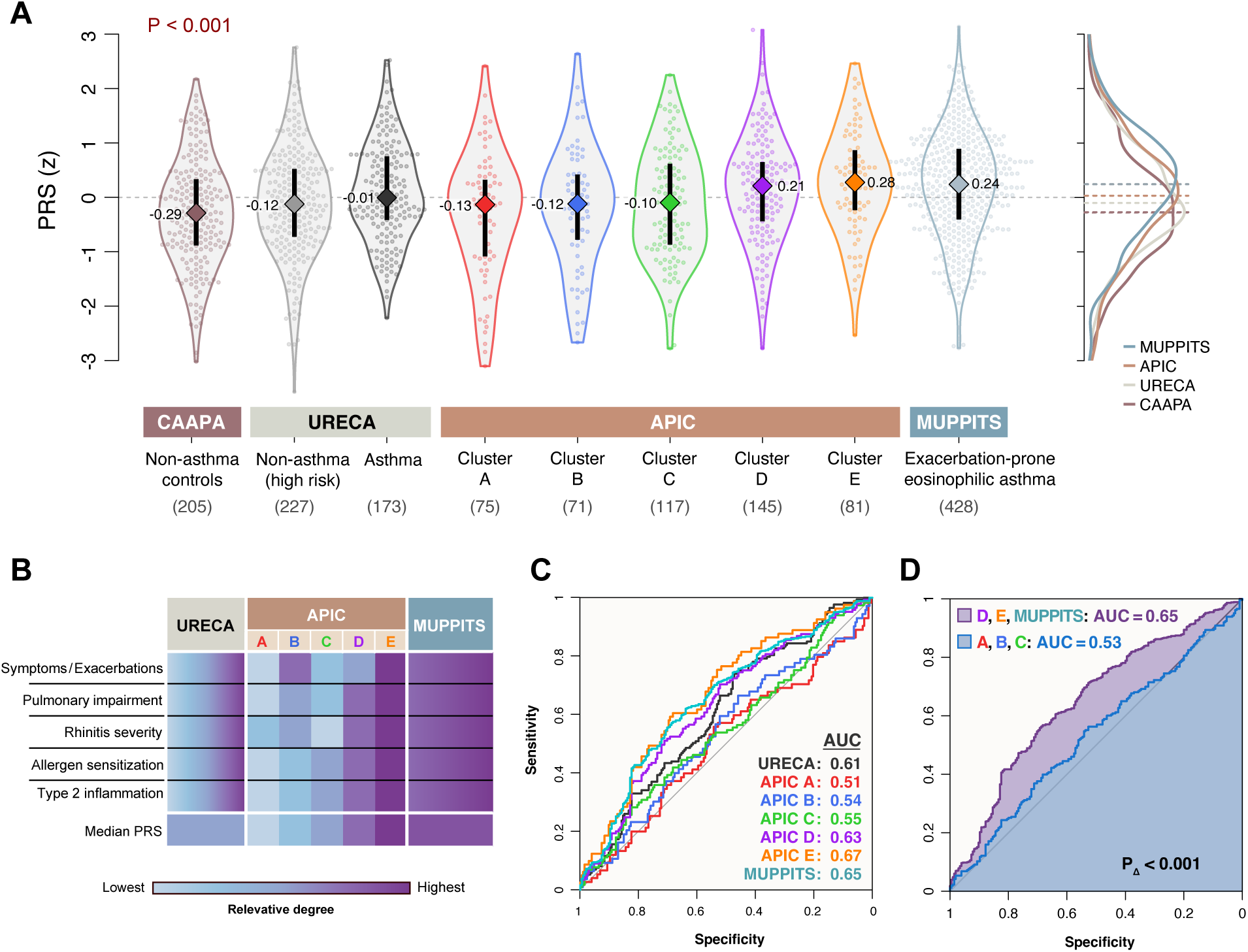
Genetic asthma prediction is stronger in more symptomatic phenotypes. A) Distributions and median values of asthma PRSs by genotyped cohort, including APIC’s previously defined sub-phenotypes (clusters A-E)^13^. B) Schematic heatmap showing the relative degrees of asthma severity for each cohort. C) Receiver operating characteristic (ROC) curves with areas under the curve (AUC) for predicting asthma using the PRS for each cohort. D) Prediction of asthma in the combined moderate-to-severe asthma cohorts (APIC D, E & MUPPITS) was significantly more accurate than for the milder asthma subphenotypes (APIC A, B, C), combined.

To more directly test whether polygenic asthma risk correlates with disease severity, we tested PRS associations with cohort-specific severity metrics. In APIC, PRSs were significantly higher in difficult-to-control vs. easy-to-control asthma^12^ (P=0.02; **Figure 3A**), indicating that children with more genetic risk alleles tended to require higher treatment levels to control their asthma. In the MUPPITS-2 trial, which investigated the effect of an anti-IL-5 inhibitor (mepolizumab) in children with exacerbation-prone eosinophilic asthma^8^, polygenic risk was significantly associated with higher exacerbation rates. However, the association was only observed in the placebo-treated group (P=0.03; **Figure 3B**). The correlation was not significant in the study arm treated with mepolizumab (P=0.53), suggesting that the biologic effectively reduced the asthma genetic effect on the exacerbation rate.

**Figure 3.**
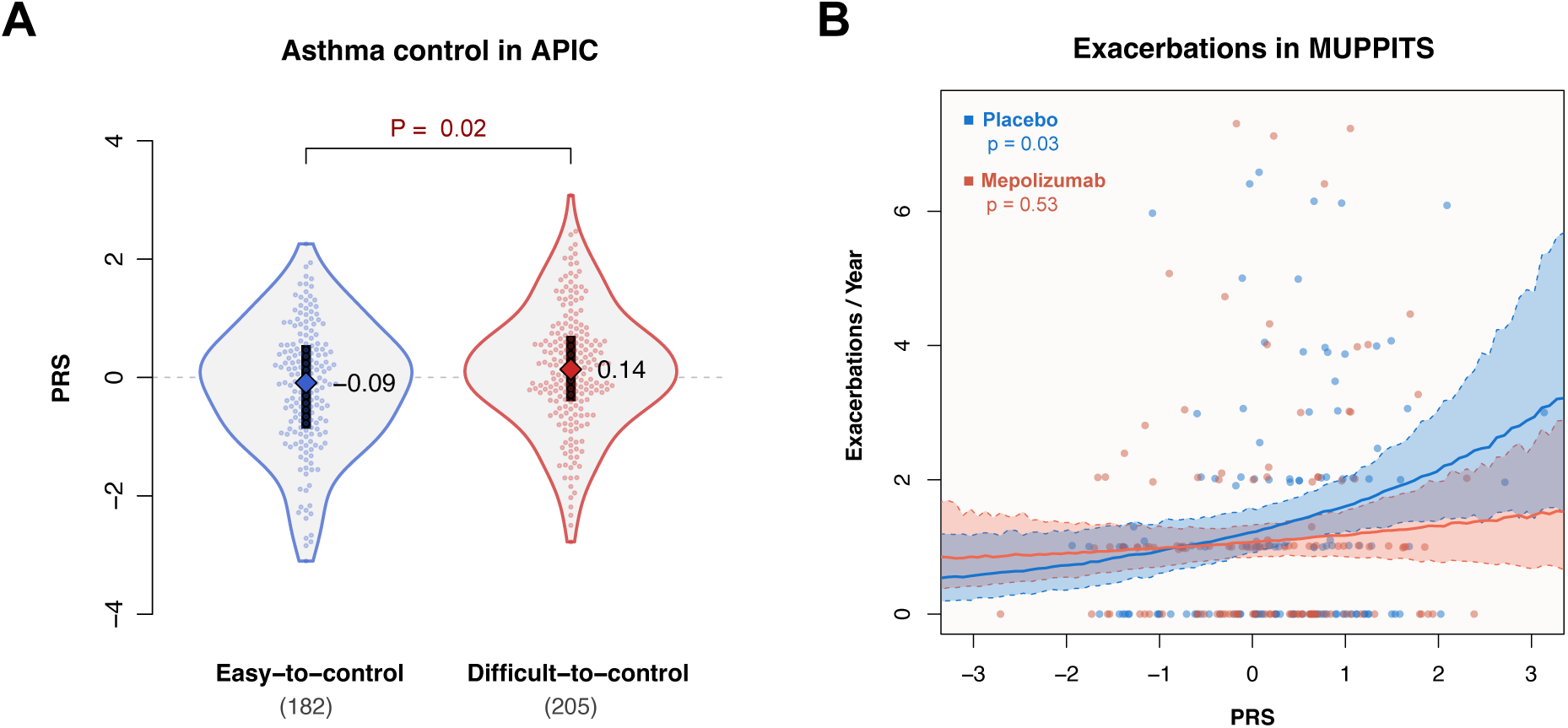
Polygenic risk is associated with asthma severity and exacerbation risk. A) Distributions and median values of asthma PRSs in APIC by required treatment step^12^ Sample sizes are shown below the x-axis labels. B) Exacerbation rate vs. PRSs in MUPPITS-2, split by treatment arm. Predicted exacerbation rates by PRS are shown for each treatment arm (solid lines) with 95% confidence intervals (shaded areas).

We next assessed associations between the PRS and quantitative biomarkers of asthma severity^18^. In children with asthma across all cohorts, the PRS was significantly correlated with higher blood eosinophil levels (P=0.01; **Figure 4A**) and lower lung function (P<0.001; **Figure 4C**), as measured by the ratio of forced expiratory volume in one second to forced vital capacity (FEV_1_/FVC). Comparing the highest to lowest PRS deciles, these effects amounted to significant median differences of 101 eosinophils per μL (P=0.02; **Figure 4B**), and -3.0% FEV_1_/FVC (P=0.005; **Figure 4D**). These findings further demonstrate that genetic asthma risk is correlated with quantitative measures relevant to asthma severity.

**Figure 4.**
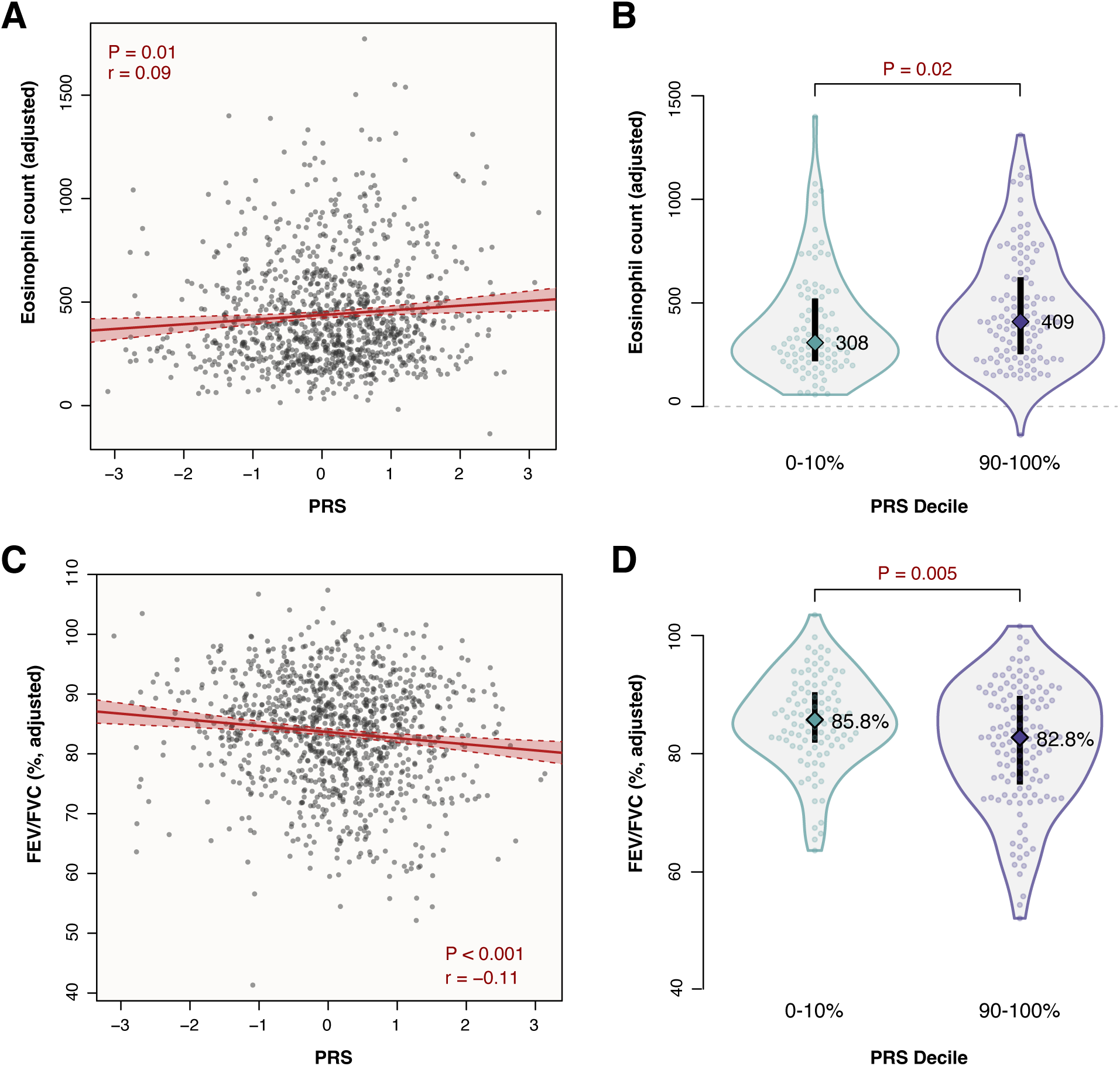
Polygenic risk is associated with quantitative biomarkers of asthma severity. A) Eosinophil count per μL, adjusted for age, sex, study, and ancestry, plotted against the PRS in a combined sample from APIC, URECA, and MUPPITS (n=1,062). The modeled correlation fit with 95% confidence interval is drawn in red. B) Distributions and median values of adjusted eosinophil counts in the lowest and highest PRS deciles. C) Adjusted FEV_1_/FVC plotted against the PRS in a combined sample from APIC, URECA, and MUPPITS (n=1,098). D) Distributions and median values of adjusted FEV_1_/FVC in the lowest and highest PRS deciles.

## DISCUSSION

Genetic disease risk depends on phenotype definitions, ancestry, and environmental exposures. The GBMI asthma GWAS that we used for modeling our PRS drew predominantly from European and East Asian adult cohorts and defined asthma according to specific International Classification of Diseases (ICD)-9 and ICD-10 codes from participant electronic health records^37^. Therefore, the asthma phenotypes, ancestries, and environments captured in the GBMI GWAS differed overall from those in the four cohorts studied here, which focused on childhood-onset asthma in urban environments in the United States, consisting mostly of Black and Hispanic populations. It has been shown that PRS predictive performance deteriorates across ancestries^38^, and we previously demonstrated how modeling genetic risk from different asthma GWASs yielded PRSs with different trait associations^7^. Furthermore, asthma cases in the GBMI GWAS consisted largely of adult-onset disease^2,7^, and the genetic architectures of childhood- and adult-onset asthma are partly distinct, with reported genetic correlations ranging from 0.67-0.78^2,39^. Although the GBMI-derived PRS was found to be the best performing asthma PRS in individuals of African ancestry^40^ and predicted asthma in this study with accuracy comparable to its performance in non-African populations^7,40^, the GBMI PRS is not a perfect proxy for genetic asthma risk in this study’s cohorts. Thus, the actual correlation between genetic asthma risk and disease severity is likely stronger than the effects we observed here.

Among patients with physician-diagnosed asthma according to ICD-10 codes, the vast majority have mild-to-moderate asthma, with reported prevalences of severe asthma ranging from 2-14%^41–44^, depending on the country and definition used for severe asthma. Asthma in the GBMI GWAS was primarily defined according to ICD codes in the constituent biobanks. Therefore, it is unlikely that the PRS correlations we observed with asthma severity are due to asthma in GBMI being disproportionately severe. Instead, our results indicated that the cumulative burden of genetic risk variants for asthma in general contributes to disease severity. This conclusion is supported by GWASs of moderate-to-severe asthma, which found that 88% of association signals that reached genome-wide significance had been previously reported in studies that predominantly assessed samples from patients with mild asthma^45,46^. Our findings expand on those from a population-based longitudinal study in European ancestry individuals, in which asthma cases with a higher polygenic burden calculated from 17 risk variants were more likely to manifest atopy, airway hyperresponsiveness, and incompletely reversible airflow obstruction, and more often experienced hospitalizations for breathing problems^47^. Our results also align with another study that found polygenic risk for asthma was significantly associated with asthma exacerbations in European ancestry individuals^48^. Similar observations, where cumulative genetic risk from low-effect variants were associated with disease severity, have been reported in other complex diseases, as well, including cardiovascular disease^49^ and autoimmune disease^50^. Such genetic effects may combine with effects from rare variants^45,51^ and environmental exposures^52^ to contribute to disease severity through shared or independent pathways^9^.

In summary, the results of our study indicate that the cumulative effect of asthma-associated genetic variants not only increase disease risk but also contribute to disease severity. By applying a GBMI-trained PRS to four multi-ancestry childhood-onset asthma cohorts from low-income urban environments, we demonstrate that the effects observed in this study transcend ancestry, asthma phenotypes, severity metrics, and environments. Genetic risk models that are more closely aligned to demographic and clinical profiles of study populations would likely more precisely capture genetic influences on severity. Future studies should consider incorporating genetic risk together with other predictive factors for developing more comprehensive and accurate prediction models of asthma severity to inform strategies for clinical prevention, intervention, and management.

## Abbreviations

APIC: Asthma Phenotypes in Inner-City Children
AUC: area under the curve
CAAPA: Consortium on Asthma in African-ancestry Populations in the Americas
FEV_1_: Forced expiratory volume in one second
FVC: Forced vital capacity
GBMI: Global Biobank Meta-analysis Initiative
GWAS: Genome-wide association study
ICD: International Classification of Diseases
IQR: interquartile range
MUPPITS: Mechanisms Underlying Asthma Exacerbations Prevented and Persistent with Immune Based Therapy: A Systems Approach
PRS: Polygenic risk score
ROC: receiver operating characteristic
URECA: Urban Environment and Childhood Asthma

## ACKNOWLEDGEMENTS

URECA V has been funded in whole or in part with Federal funds from the National Institute of Allergy and Infectious Diseases, National Institutes of Health, and Department of Health and Human Services under Cooperative Agreement number 1UM1AI160040, HHSN272200900052C, HHSN272201000052I, 1UM1AI114271-01, and UM2AI117870. Additional support was provided by the National Center for Research Resources, National Center for Advancing Translational Sciences, and National Institutes of Health, under grants NCRR/NIHRR00052, NCRR/NIHM01RR00533, NCRR/NIH1UL1RR025771, NCRR/NIHM01RR00071, 1UL1RR024156, NCRR/NIH RR00052, UL1TR001079, NCRR/NIHM01RR00533, NCRR/NIH1UL1RR025771, NCRR/NIHM01RR00071, NCRR/NIH1UL1RR024156, NCATS/NIHUL1TR000040, 5UL1RR024992-02, NCATS/NIHUL1TR001079, NCATS/NIH1UL1TR001430, NCATS/NIHUL1TR001873, and NCATS/NIH UL1TR002345.

The APIC project has been funded in whole or in part with Federal funds from the National Institute of Allergy and Infectious Diseases, National Institutes of Health, and Department of Health and Human Services under Contract numbers HHSN272200900052C, HHSN272201000052I, 1UM1AI114271-01, and UM2AI117870. Additional support was provided by the National Center for Research Resources, and National Center for Advancing Translational Sciences, National Institutes of Health, under grants NCATS/NIH UL1TR000150, NCRR/NIH UL1TR000451, UL1TR001105, NCRR/NIH 1UL1RR025780 (CCTSI is supported in part by Colorado CTSA Grant UL1TR000154 from NCATS/NIH UL1TR001082), NCATS/NIH UL1TR000040, NCATS/NIH UL1TR000075, NCRR/NCAT/NIH UL1TR000077-04. GlaxoSmithKline (GSK) provided Ventolin HFA, Flonase, Flovent 50 mcg, Flovent 100 mcg, Flovent 250 mcg , Advair 250/50 mcg, Advair 500/50 mcg under a clinical trial agreement with NIH NIAID. None of these companies had a role in the development or approval of the protocol, conduct of the trial, data analysis, manuscript preparation, or the decision to submit for publication.

The MUPPITS-1 and MUPPITS-2 projects have been funded in whole or in part with Federal funds from the National Institute of Allergy and Infectious Diseases, National Institutes of Health, and Department of Health and Human Services under Contract numbers 1UM1AI114271-01 and UM2AI117870. Additional support was provided by the National Center for Research Resources, National Center for Advancing Translational Sciences, and National Institutes of Health, under grants UL1TRG01422 (05/19/08 – 04/29/12), CTSA Grant UL1RR025741 (04/30/12-08/11/15), CTSA Grant UL1TR000150 (08/12/15-03/31/19), CTSA Grant UL1TR001422, NIH/NCATS Colorado CTSA UL1 TR002535, NCATS/NIH UL1TR001876, and NIH/CTSA 5UL1TR001425-03.

Dr. Becker’s and Gergen’s co-authorship of this report does not necessarily represent the views of the National Institute of Allergy and Infectious Diseases, the National Institutes of Health or any other agency of the United States Government.

## DATA AVAILABILITY

Phenotype, genotype, GWAS summary statistics, and whole-genome sequencing files are available in dbGaP under accession phs002921 for the APIC and URECA studies and under accession phs004273 for the MUPPITS studies.

